# Ventricular and Atrial Pressure-Volume Loops: Analysis of the Effects Induced by Right Centrifugal Pump Assistance

**DOI:** 10.1101/2022.03.22.22272760

**Authors:** Beatrice De Lazzari, Attilio Iacovoni, Massimo Capoccia, Silvia Papa, Roberto Badagliacca, Carmine Dario Vizza, Claudio De Lazzari

## Abstract

**Background and Objective:** The main indications for right ventricular assist device (RVAD) support are right heart failure after implantation of a left ventricular assist device (LVAD) or early graft failure following heart transplantation. About 30-40% of patients will need RVAD support after LVAD implantation. Pulmonary hypertension is also an indication for right heart assistance. Several types of RVAD generating pulsatile or continuous flow are available on the market. These assist devices can be connected to the cardiovascular system in different ways. We sought to analyse the effects induced by different RVAD connections when right ventricular elastance is modified using a numerical simulator. The analysis was based on the behaviour of both left and right ventricular and atrial loops in the pressure-volume plane.

**Methods:** New modules of the cardiovascular network and a right ventricular centrifugal pump were implemented in CARDIOSIM^©^ software simulator platform. The numerical pump model generated continuous flow when connected in series or parallel to the right ventricle. When the RVAD was connected in series (parallel), the pump removed blood from the right ventricle (atrium) and ejected it into the pulmonary artery. In our study, we analysed the effects induced by RVAD support on left/right ventricular/atrial loops when right ventricular elastance slope (Ees_RIGHT_) changed from 0.3 to 0.8 mmHg/ml with the pump connected either in series or parallel. The effect of low and high rotational pump speed was also addressed.

**Results:** Percentage changes up to 60% were observed for left ventricular pressure-volume area and external work during in-parallel RVAD support at 4000 rpm with Ees_RIGHT_ = 0.3mmHg/ml. The same pump setting and connection type led to percentage variation up to 20% for left ventricular ESV and up to 25% for left ventricular EDV with Ees_RIGHT_ = 0.3mmHg/ml. Again the same pump setting and connection generated up to 50% change in left atrial pressure-volume loop area (PVLA_L-A_) and only 3% change in right atrial pressure-volume loop area (PVLA_R-A_) when Ees_RIGHT_ = 0.3mmHg/ml. Percentage variation was lower when Ees_RIGHT_ was increased up to 0.8 mmHg/ml.

**Conclusion:** Early recognition of right ventricular failure followed by aggressive treatment is desirable to achieve a more favourable outcome. RVAD support remains an option for advanced right ventricular failure although onset of major adverse events may preclude its use.

## INTRODUCTION

The main indications for right ventricular assist device (RVAD) support are right heart failure after implantation of a left ventricular assist device (LVAD) or early graft failure following heart transplantation. Measurement of right heart function is often overlooked during daily clinical practice. The study by [1] is an attempt to better understand RV pathophysiology in the setting of pulmonary hypertension, which represents the prevalent pathological condition for RVAD indication, whether to support a failing left ventricle or a primarily involved RV [2].

However, the lack of data does not allow an in-depth analysis of right ventricular and atrial behaviour during RVAD support. An attempt in this direction can be made using a numerical simulator of the cardiovascular system with a view to reproduce pathological conditions requiring right ventricular assistance.

The aim of our study was the analysis of the haemodynamic and energetic variables of both ventricles and atria in different cardiovascular conditions reproduced by changing right ventricular contractility from 0.3 to 0.8 mmHg/ml during RVAD support with different rotational speeds.

The first step of this work required the implementation of two new modules within CARDIOSIM^©^ software simulation platform, which would reproduce the characteristics of the left circulatory network and a continuous flow centrifugal pump (RVAD) to be connected either in series or parallel to the right ventricle. The new modules were based on a 0-D (lumped-parameter) numerical model including input and output cannula of the RVAD.

We simulated RVAD support following in series and parallel connection driven by different rotational speeds in a heart failure setting. Subsequently, the right ventricular elastance was increased from 0.3 to 0.8 mmHg/ml in a stepwise manner. During each setting, we focused our attention on the following hemodynamic and energetic variables:

✓ Right and left ventricular end-systolic volume (ESV_R-V_ and ESV_L-V_);
✓ Right and left ventricular end-diastolic volume (EDV_R-V_ and EDV_L-V_);
✓ Stroke volume (SV);
✓ Right and left ventricular external work (EW_R-V_ and EW_L-V_) and pressure-volume area (PVA_R-V_ and PVA_L-V_);
✓ Right and left atrial end-systolic volume (ESV_R-A_ and ESV_L-A_);
✓ Right and left atrial end-diastolic volume (EDV_R-A_ and EDV_L-A_);
✓ Right and left atrial pressure-volume loop area (PVLA_R-A_ and PVLA_L-A_);
✓ Cardiac output (CO);
✓ Systolic, diastolic and mean systemic aortic pressure (AoP);
✓ Systolic, diastolic and mean pulmonary arterial pressure (PAP);
✓ Pulmonary capillary wedge pressure (PCWP);
✓ Right and left atrial pressure (RAP and LAP).

## MATERIAL AND METHODS

### The heart and circulatory numerical network

CARDIOSIM^©^ software platform has been previously described [3]-[8]. The modules of the software simulator are: systemic and pulmonary arterial section, systemic and pulmonary venous section and the coronary circulation. Native left and right ventricles, atria and septum reproduce the entire cardiac activity; they are implemented in a single module (Fig. 1) using the time-varying elastance concept [4],[5]. The ventricular, atrial and septal activity is synchronized with the electrocardiographic (ECG) signal [5]. Using the modified time-varying elastance theory, the inter-ventricular septum (IVS) interaction and the instantaneous left and right ventricular pressure can be reproduced by [5],[10]:

**Fig. 1.**
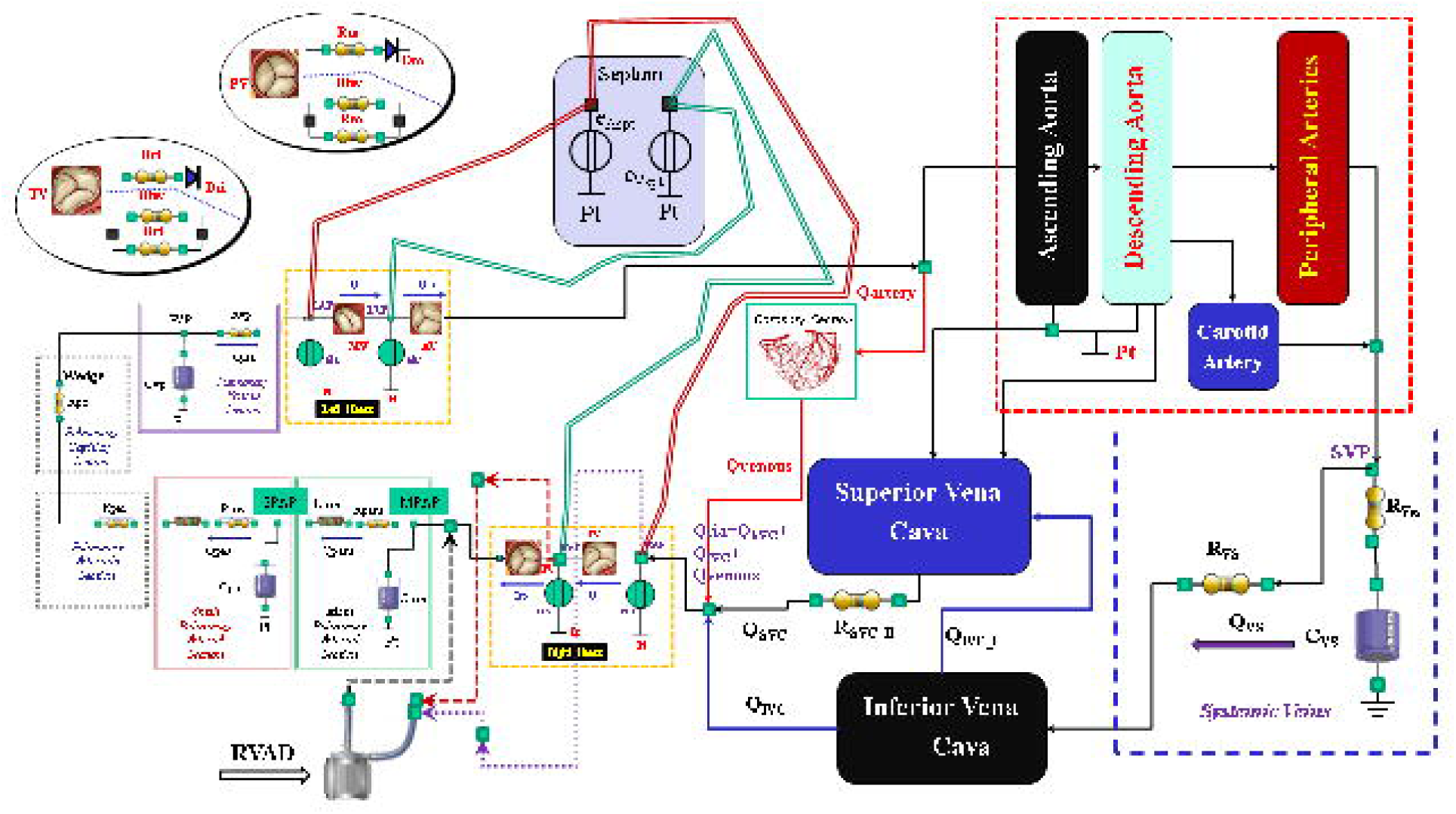
Electric analogue of the cardiovascular system. The network is assembled with septum, left and right heart, main and small pulmonary arterial sections, pulmonary arteriole and capillary sections, pulmonary venous section. The left circulation include ascending and descending aorta compartments, peripheral arteries and carotid artery sections, coronary circulation, superior and inferior vena cava sections and systemic veins compartment. RVAD is the right ventricular assist device. Table 2 lists the symbols used.

**Fig. 1a.**
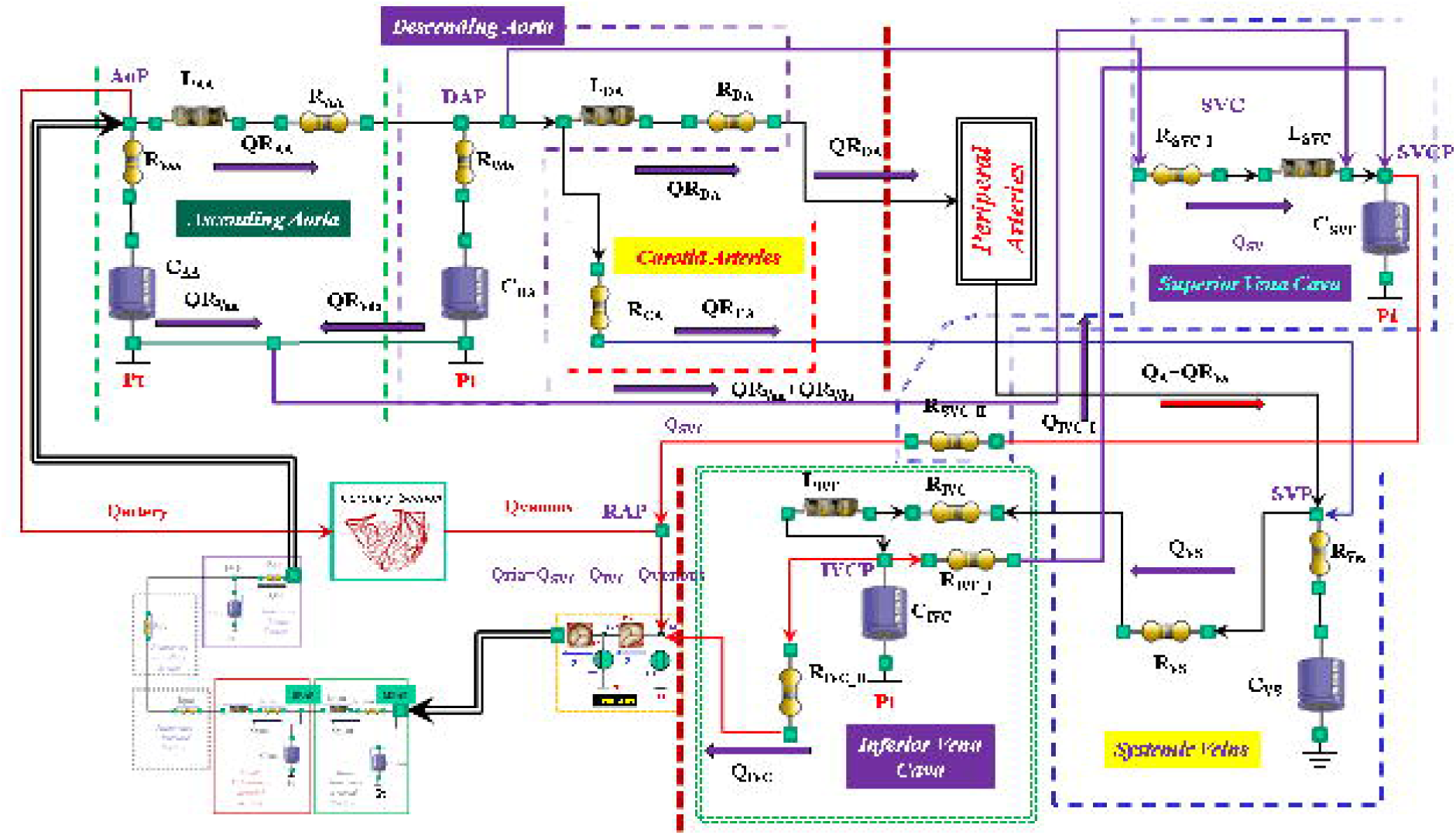
The behavior of the ascending aorta is simulated with resistances R_AA_ and R_Vaa_, inertance L_AA_ and compliance C_AA_. QR_AA_ is the flow through the resistance and inertance. The descending aorta is implemented with resistances R_DA_ and R_Vda_, inertance L_DA_ and compliance C_DA_. QR_DA_ is the flow through the resistance (R_DA_) and inertance (L_DA_). The carotid arteries section is reproduced with a simple resistance (R_CA_). The superior vena cava module consists of resistances R_SVC_I_ and R_SVC_II_, inertance L_SCV_ and compliance C_SVC_. The inferior vena cava module is modelled with resistances R_IVC_, R_IVC_I_ and _RIVC_II_, inertance L_IVC_ and compliance C_IVC_. The intrathoracic pressure (Pt) affects compliances C_AA_, C_DA_, C_IVC_ and C_SVC_. Table 2 lists the symbols used.

**Fig. 1b.**
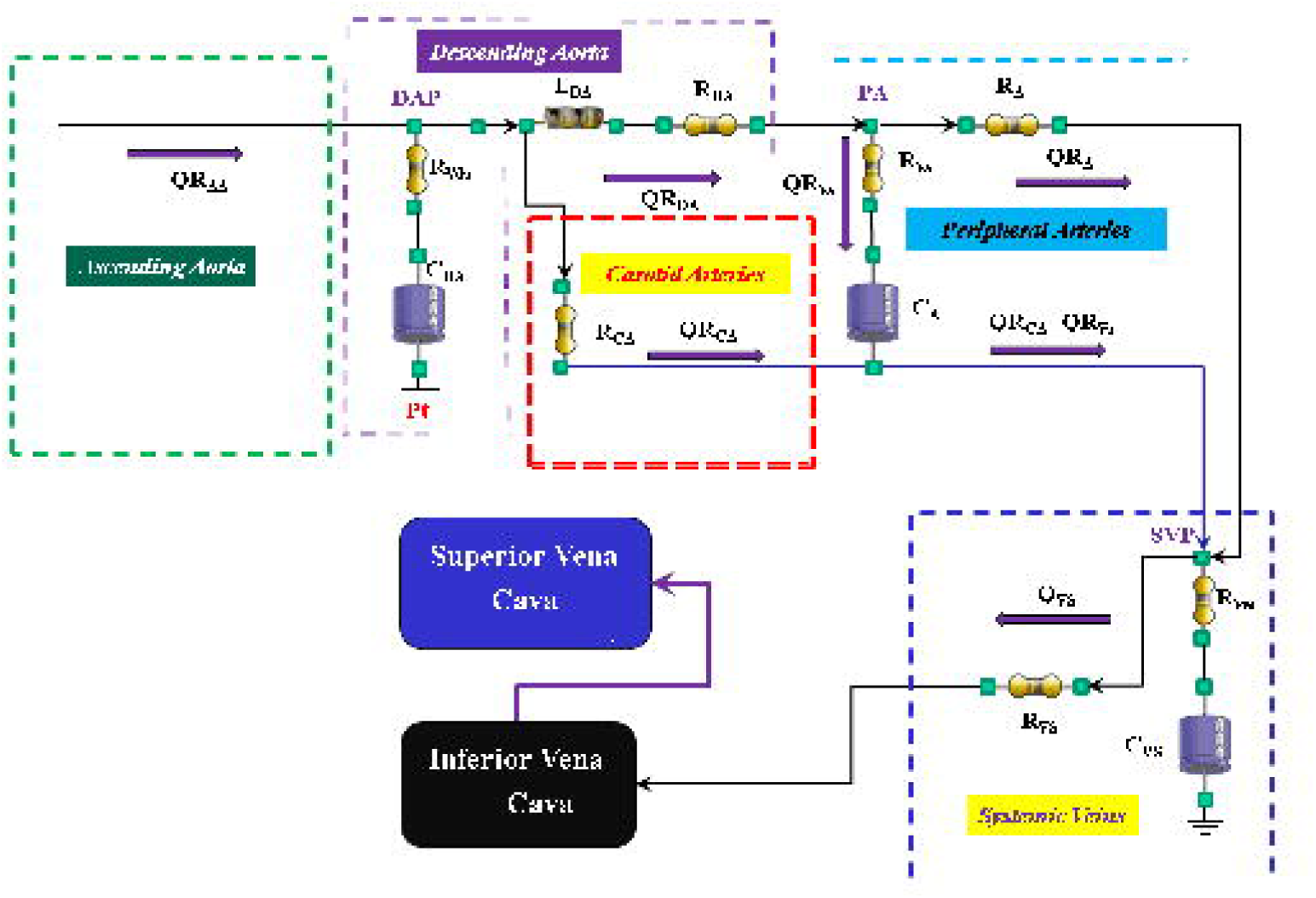
The peripheral arteries module is modelled with resistances R_A_ and R_Va_ and compliance C_A_. The resistor R_Va_ account for viscous losses of the vessels wall. QR_A_ is the blood flow outside the compartment; it is a part of the blood that reaches the systemic veins compartment.

**Fig. 1c.**
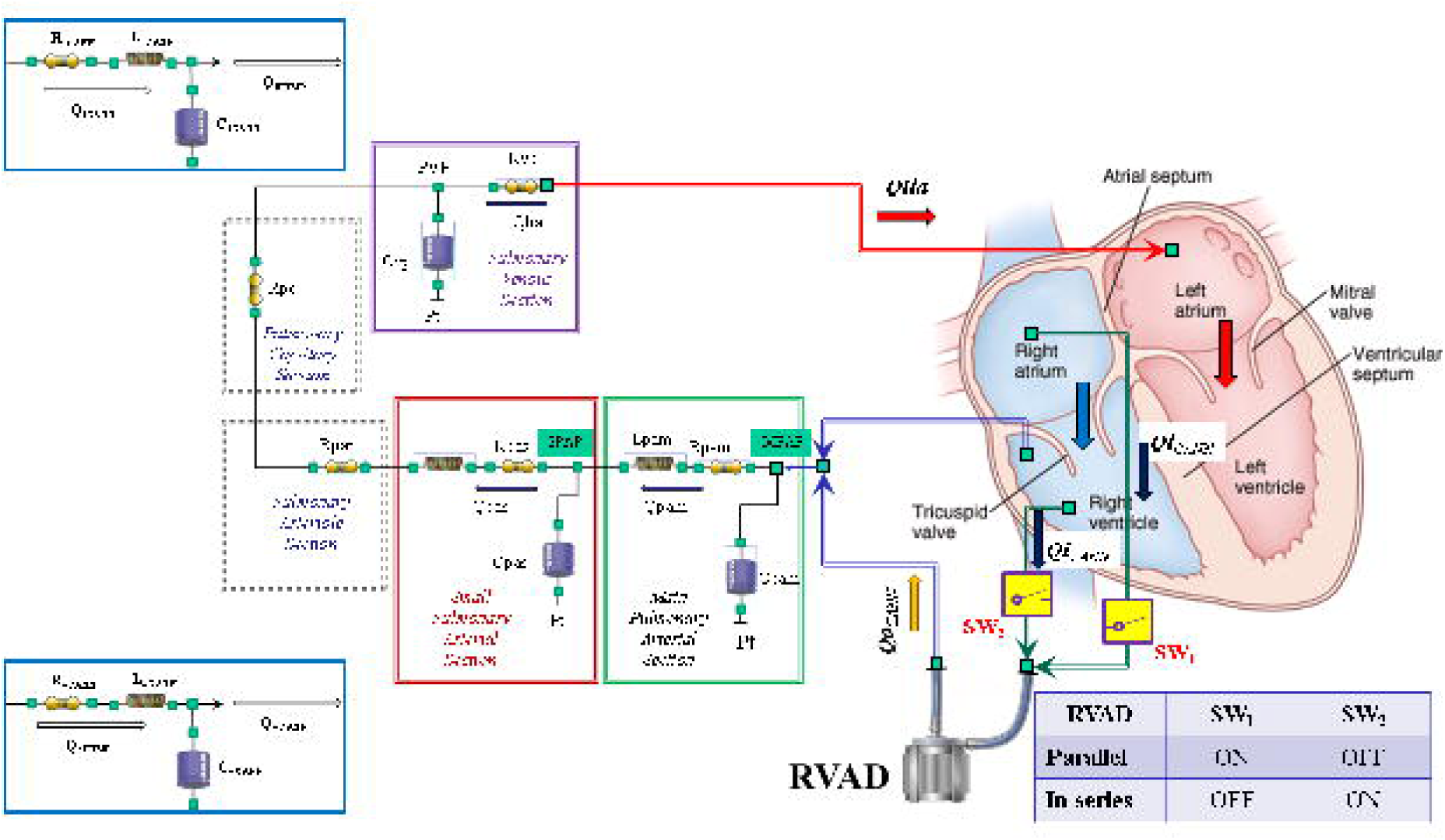
Schematic representation of RVAD connection. When the right ventricular assist device is connected in parallel, blood is removed from the right atrium (SW_1_=ON and SW_2_=OFF) and ejected into the pulmonary artery. When RVAD is connected in series, blood is removed from the right ventricle (SW_1_=OFF and SW_2_=ON) and ejected into the pulmonary artery. The input (output) RVAD cannula is modelled with RLC elements. Q_oPUMP_ (Q_iPUMP_) is the output (inlet) flow rate from the pump. Q_oCANN_ (Q_iCANN_) is the output (inlet) flow rate from the cannula. The electrical analogue of the pulmonary circulation is described in **Error! Reference source not found**..

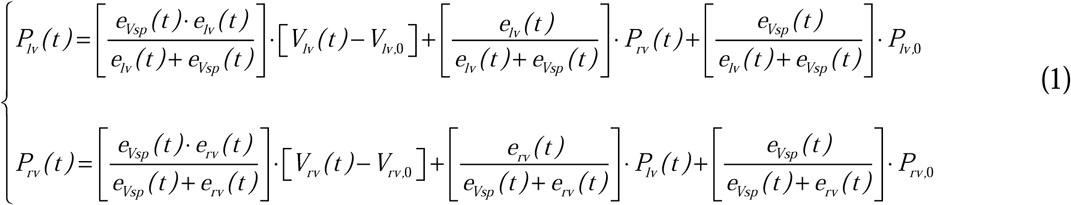

In the same way, the inter-atrial septum (IAS) interaction and the left and right instantaneous atrial pressure can be reproduced by:

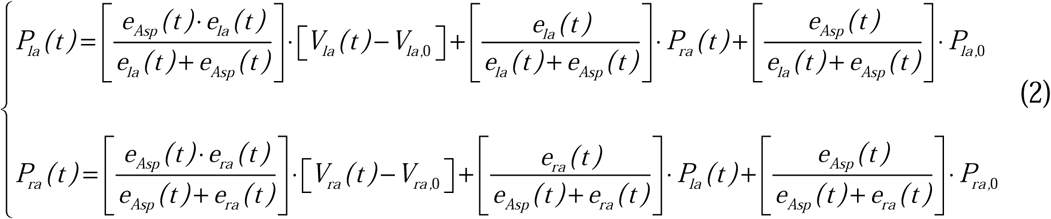

The symbols used in Eqs. (1) and (2) have been listed in Table 1. These features allow the simulation of inter-ventricular and intra-ventricular dyssynchrony [10].

Specific modules of the coronary circulation (Fig. 1) are also available in CARDIOSIM^©^ platform [3].

For the purposes of our study, we have assembled the cardiovascular network with the new module of the systemic circulation whilst the behavior of the heart is modelled as described in [3],[4]. The systemic venous section [3],[6] and the entire pulmonary circulation [7],[8],[11],[12] are modelled as described in current literature. We have selected the module presented in [13] for the coronary circulation. The tricuspid, mitral, pulmonary, and aortic valves are modelled with an ideal diode: when the pressure across the valve is positive, the valve opens and allows the flow of blood; when the pressure is less than or equal to zero, the valve is closes and the flow of blood is zero [3]-[5],[14]

### New lumped-parameter model of the systemic circulation

The new module is described in Fig. 1 using resistance, inertance and compliance (RLC) elements. The systemic network consists of the following compartments: ascending and descending aorta, carotid artery, peripheral arteries and superior and inferior vena cava (Fig. 1a).

The ascending (descending) aorta is modelled using two resistances R_AA_ and R_Vaa_ (R_DA_ and R_Vda_), inertance L_AA_ (L_DA_) and compliance C_AA_ (C_DA_). A single resistance (R_CA_) reproduces the behavior of the carotid district. The peripheral arterial circulation is reproduced with resistances R_A_ and R_Va_ and with compliance C_A_ (Fig. 1b). The superior vena cava compartment consists of resistances R_SVC_I_ and R_SVC_II_, inertance L_SVC_ and compliance C_SVC_. The systemic venous network is implemented using compliance C_VS_ and resistances R_VS_ and R_Vvs_ (Fig. 1a). Finally, the inferior vena cava district is modelled with resistances R_IVC_, R_IVC_I_ and R_IVC_II_, inertance L_IVC_ and compliance C_IVC_ (Fig. 1a). The resistances R_Vaa_, R_Vda_, R_Va_, and R_Vvs_ account for viscous losses of the vessel wall. Pt is the mean intrathoracic pressure.

### Right ventricular assist device (RVAD)

A 0-D numerical model described in [7] was used to implement a centrifugal pump that reproduced the behavior of the right ventricular assist device. The pump can be connected in series or parallel to the right ventricle with two cannulae modelled using RLC elements (Fig. 1c). When the RVAD removes blood from the right atrium (parallel connection – SW1=ON and SW2=OFF in Fig. 1c), the flow through the inlet cannula is:

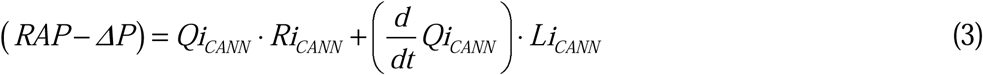

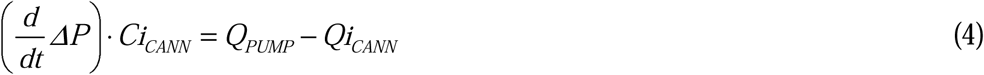

*RAP* is the right atrial pressure, *Li*_*CANN*,_ *Ri*_*CANN*_ and *Ci*_*CANN*_ are the inertance, resistance and compliance of the inlet cannula, respectively (Fig 1c). *Qi*_*CANN*_ (*Q*_*PUMP*_) is the flow through the inlet cannula (generated by the centrifugal pump). *ΔP* is the pressure on the pump head.

When the RVAD removes blood from the right ventricle (in series connection - SW1=OFF and SW2=ON in Fig. 1c), Eq. 3 becomes:

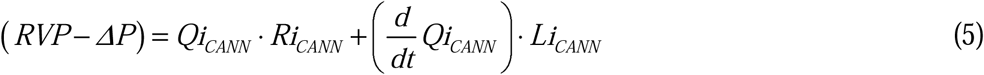

*RVP* is the right ventricular pressure.

When the RVAD ejects blood, the flow through the outlet cannula is:

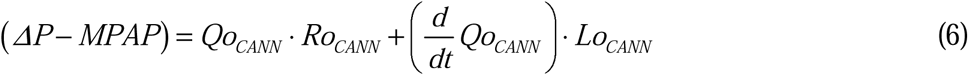

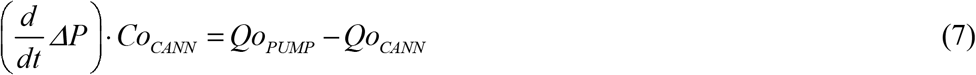

*MPAP* is the mean pulmonary artery pressure (Fig. 1c), *Ro*_*CANN*_ and *Co*_*CANN*_ are the inertance, resistance and compliance of the outlet cannula, respectively. *Qo*_*CANN*_ is the flow through the outlet cannula.

### Simulation Protocol

For the purposes of our simulations, we considered values for right and left ventricular elastance that could reproduce a realistic diseased heart. According to the available literature, normal values for right ventricular elastance fluctuate around 1 mmHg/ml [15],[16] and range from 1.6 to 5 mmHg/ml for left ventricular elastance [17],[18]. Therefore, we considered Ees_LEFT_ = 0.7 mmHg/ml for the left ventricle and Ees_RIGHT_ = 0.3 mmHg/ml for the right ventricle as reference values for a failing heart. Our simulation approach consisted of three steps. In the first step after setting HR= 90 bpm, the slope of left ventricular End-Systolic Pressure-Volume Relationship (ESPVR) Ees_LEFT_=0.7 mmHg/ml and the slope of right ESPVR Ees_RIGHT_=0.3 mmHg/ml [8], the simulator generated the following values: cardiac output (CO) 4.51 l/min, aortic systolic (diastolic) pressure 82.1 (60.4) mmHg, mean right atrial pressure 23.3 mmHg, pulmonary systolic (diastolic) pressure 51.2 (31.6) mmHg and pulmonary capillary wedge pressure 20.7 mmHg. The mean pressure (flow) value was calculated as the mean value of all blood pressure (flow) measurements during a cardiac cycle.

In the second step, the slope of right ESPVR Ees_RIGHT_=0.3 mmHg/ml was set to 0.4, 0.5, 0.6, 0.7 and 0.8 mmHg/ml [1],[19] and for each value the parameters described above were measured.

In the third step, RVAD support was applied both in series and parallel mode with rotational speed of 2000, 2500, 3000, 3500 and 4000 rpm. In the fourth step, the slope of right ESPVR was changed from 0.3 to 0.8 mmHg/ml (0.1 mmHg/ml stepwise increase) during RVAD support connected in series and parallel mode. The measured parameters were those described above.

Considering that we did not measure data in patients undergoing RVAD support to allow us to reproduce their haemodynamic conditions with our numerical simulator, and given that literature data are largely incomplete, we decided against a direct data comparison. Therefore, we considered percentage variation to evaluate the trend of the effects induced by RVAD support on the haemodynamic and energetic variables in line with other available simulation work.

## RESULTS

Figure 2 shows the effects induced by different values for ESPVR slope (Ees_RIGHT_) on left and right atrial pressure-volume loop area (left upper panel). The percentage changes calculated with respect to the reference value for Ees_RIGHT_=0.3 mmHg/ml have been listed. The red bars show the percentage change with respect to the reference value for PVLA_L-A_ increase at high Ees_RIGHT_ values. On the contrary, the PVLA_R-A_ (yellow bars) decreases when the percentage change is referred to high Ees_RIGHT_ values. These results show that when Ees_RIGHT_ increases from 0.3 to 08 mmHg/ml, the area of the pressure-volume loop of the right atrium decreases whilst the area of the pressure-volume loop of the left atrium increases (Fig. 3). The left and right lower panel shows the left and right atrial pressure-volume loop obtained with different Ees_RIGHT_ values. The black pressure-volume loop was obtained setting Ees_RIGHT_=0.3 mmHg/ml, while the blue pressure-volume loop was obtained setting Ees_RIGHT_=0.6 mmHg/ml. The left (left side) and the right (right side) ventricular pressure-volume loops are placed in the upper panels. When Ees_RIGHT_ increases, EW_L_V_ and PVA_L-V_ (left upper panel) increase leading to right-sided shift with increased left ESV and EDV. The effect induced by different Ees_RIGHT_ values on left and right ventricular EW (EW_L-V_ and EW_R-V_) are reported in Fig. 2 (left lower panel). The effect induced on EDV and ESV of the left (right) atrium is reported in the upper (lower) right panel (Fig. 2). The effect induced on right atrial ESV is more evident than the one produced on right atrial EDV. Figure 3 (right upper panel) shows that an increase in Ees_RIGHT_ leads to an increase in right ventricular external work and PVA_R-V_ with left-sided shift and decrease of both right ESV and EDV. Figure 4 shows the effect induced by RVAD support driven in parallel connection (left panels) on left ventricular PVA_L-V_ (left upper panel) and EW_L-V_ (left lower panel). The effect induced on PVA_L-V_ and EW_L-V_ when the RVAD is driven in series is available in the right panels. The data were measured for different values of Ees_RIGHT_ (0.3; 0.5 and 0.8 mmHg/ml) and with increasing RVAD rotational speed (2000; 2500; 3000; 3500 and 4000 rpm). For each rotational speed and Ees_RIGHT_ values, the percentage changes calculated with respect to the reference value measured in pathological conditions have been listed (Fig. 4). The highest percentage changes in PVA_L-V_ and EW_L-V_ (50% or more for both variables) were recorded in a diseased condition with right ventricular elastance set to Ees_RIGHT_=0.3 mmHg/ml during in-parallel RVAD assistance with pump rotational speed at 4000 rpm. Figure 5 shows the effect induced by in-parallel RVAD support on left ventricular ESV_L-V_ (left upper panel) and EDV_L-V_ (left lower panel). The effect induced on ESV_L-V_ (right upper panel) and EDV_L-V_ (right lower panel) by in-series RVAD support is also available for comparison purposes. When the rotational speed of the pump was set to 2000 rpm, the percentage changes for each value of right ventricular ESPVR slope were no more than 3%. The effects induced by different rotational speed of the RVAD on left and right atrial pressure-volume loop area are available in Fig. 6 for three different values of right ESPVR slope. In addition, for each value of the slope, the effect induced by the pump has been calculated in percentage terms with respect to the value obtained without assistance. In the case of the right atrium, we observed a higher percentage reduction of the loop area (from 2% to 9%) during in-series assistance (left lower panel). In the case of parallel connection (left upper panel), the percentage reduction of the pressure-volume loop area of the right atrium becomes more evident at high values for both the slope and the pump rotational speed. Right heart assistance produces significant effects on the left atrium loop area (right panels). Percentage variations between 20% and 50% are observed when the RVAD is connected in parallel to the right ventricle (right upper panel). Whether in-series or in-parallel assistance is considered, the most significant percentage variations are observed when the right ventricular ESPVR slope is set to Ees_RIGHT_=0.3 mmHg/ml. The effects induced by RVAD support on the right ventricular flow output and on the total cardiac output are showed in Fig. 7. RVAD parallel assistance generated a lower reduction in right ventricular output compared to in series assistance for each Ees_RIGHT_ value and each pump rotational speed. More specifically, when the pump speed was set to 4000 rpm, parallel assistance induced a reduction between 20% and 45% compared to the baseline value while the in series assistance induced a reduction between 38% and 78%. The upper panels in Fig. 7 show that parallel RVAD assistance generates a higher increase in total cardiac output compared to in series assistance at pump speeds higher than 3500 rpm (for each Ees_RIGHT_ value).

**Fig. 2.**
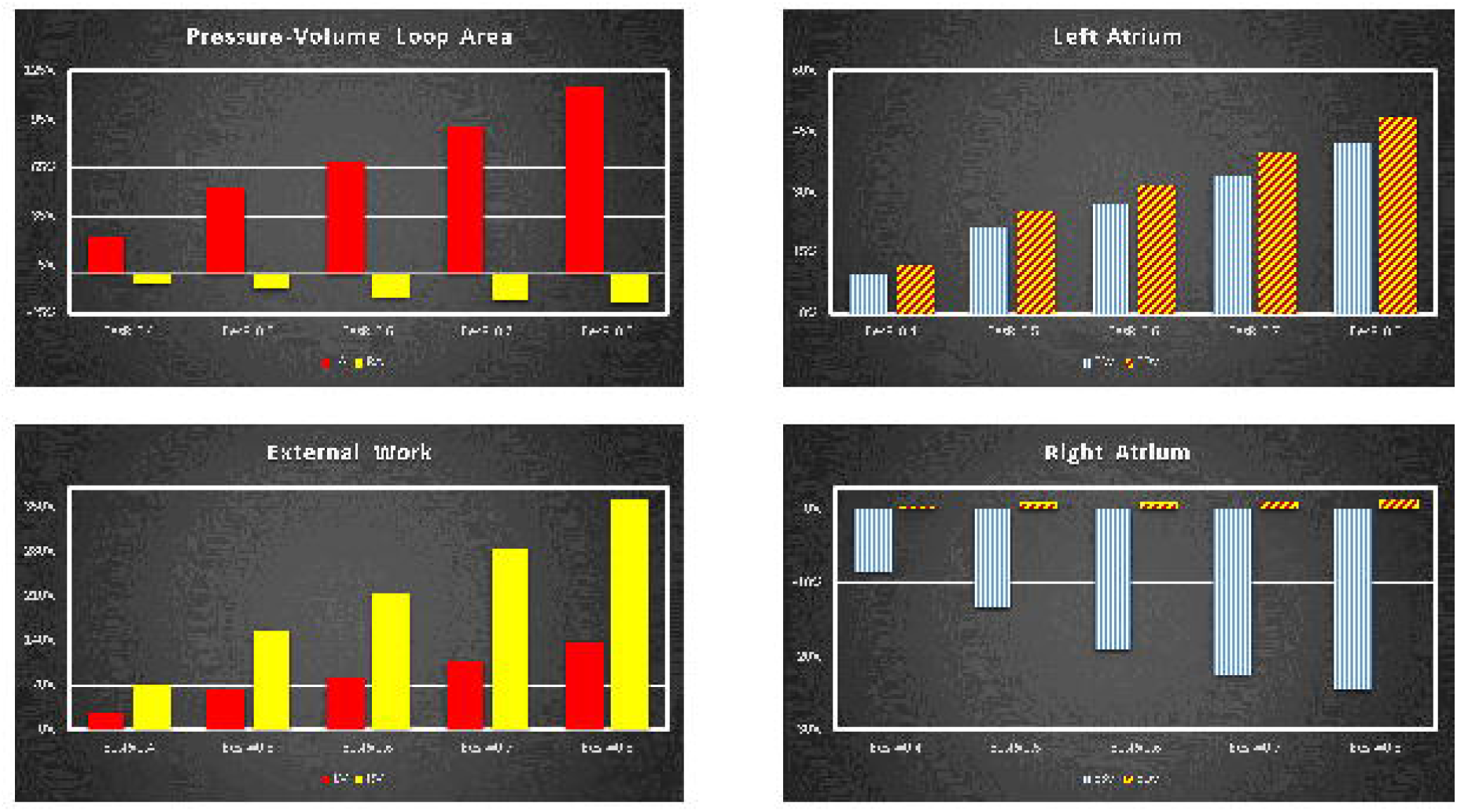
Relative changes calculated in comparison to Ees_RIGHT_=0.3 mmHg/ml for different Ees_RIGHT_ values (0.4-0.8 mmHg/ml). The relative changes of the pressure-volume loop area of the left and right atrium (external work of the left and right ventricle) are reported in the left upper (lower) panel. The right upper (lower) panel shows the relative changes of left (right) atrial end systolic and end diastolic volume.

**Fig. 3.**
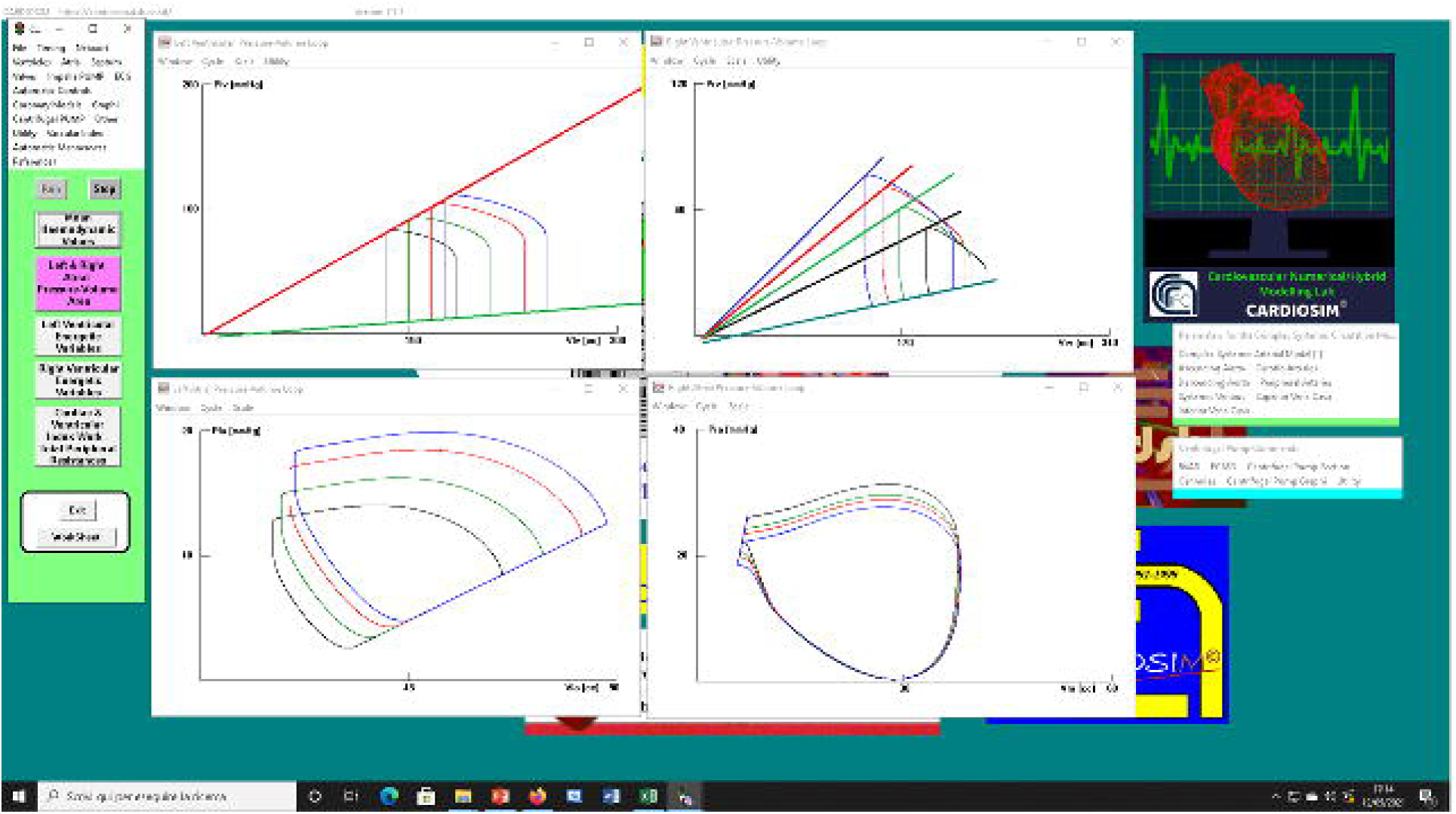
Screen output from CARDIOSIM^©^ software simulator. The left (right) upper panel shows four left (right) ventricular pressure-volume loops obtained setting EesR_lrl_Ees_RIGHT_=0.3 mmHg/ml (black line), EesR_lrl_Ees_RIGHT_=0.4 mmHg/ml (green line), EesR_lrl_Ees_RIGHT_=0.5 mmHg/ml (red line) and EesR_lrl_Ees_RIGHT_=0.6 mmHg/ml (blue line) respectively. The left (right) lower panel shows four left (right) atrial pressure-volume loops obtained changing the slope of the right ventricular elastance as previously described.

**Fig. 4.**
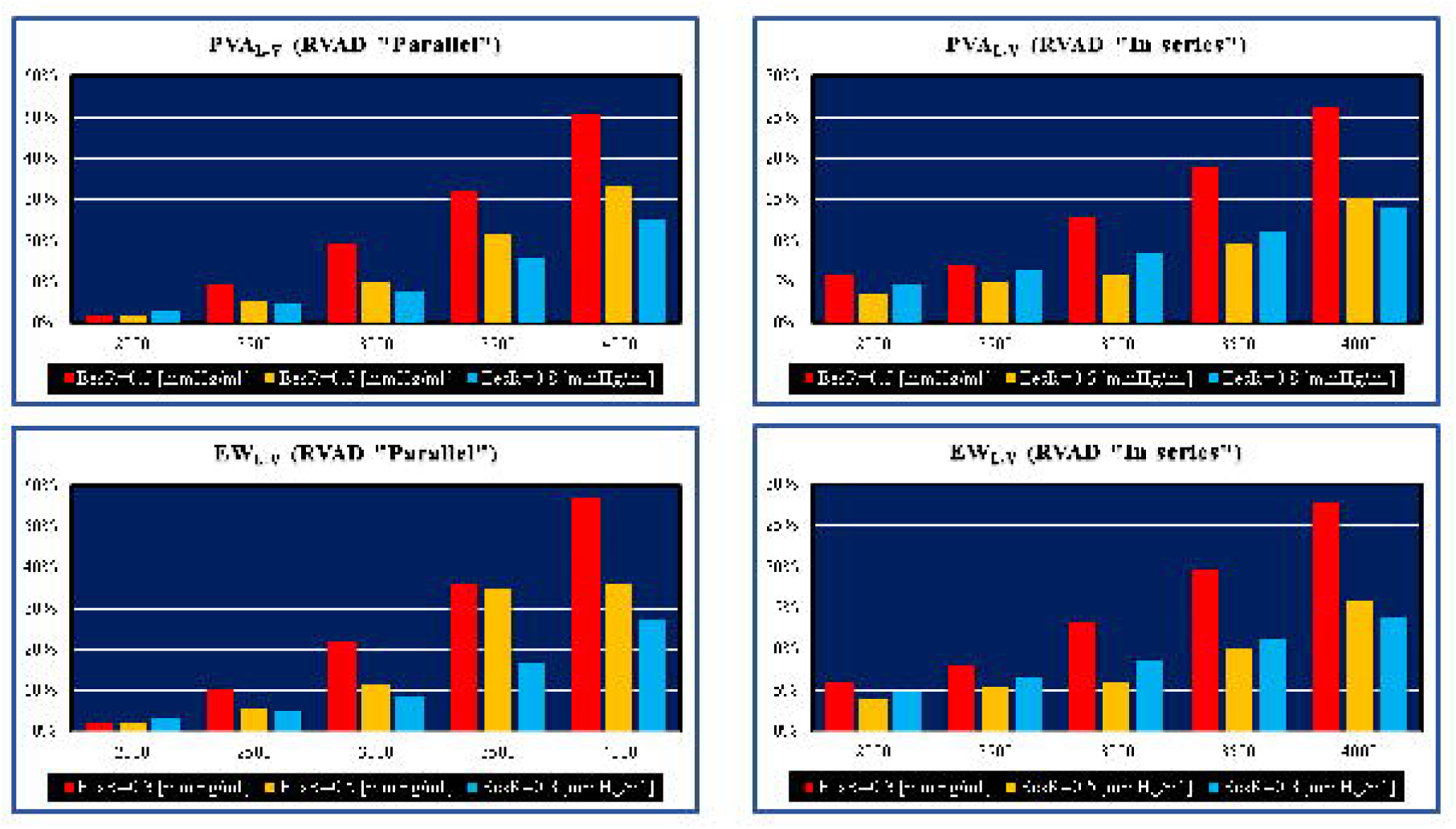
Relative changes calculated in comparison to pathological conditions (EesR_lrl_Ees_RIGHT_=0.3; 0.5 and 0.8 mmHg/ml) for different type of RVAD connection and different rotational speed. For each values of Ees_RIGHT_, the relative change was calculated when the RVAD was connected in parallel and in series. The left upper (lower) panel shows the relative changes in the left ventricular pressure-volume area (external work) when the RVAD was connected in parallel to the right ventricle. The right upper (lower) panel shows the relative changes in the left ventricular pressure-volume area (external work) when the RVAD was connected in series to the right ventricle.

**Fig. 5.**
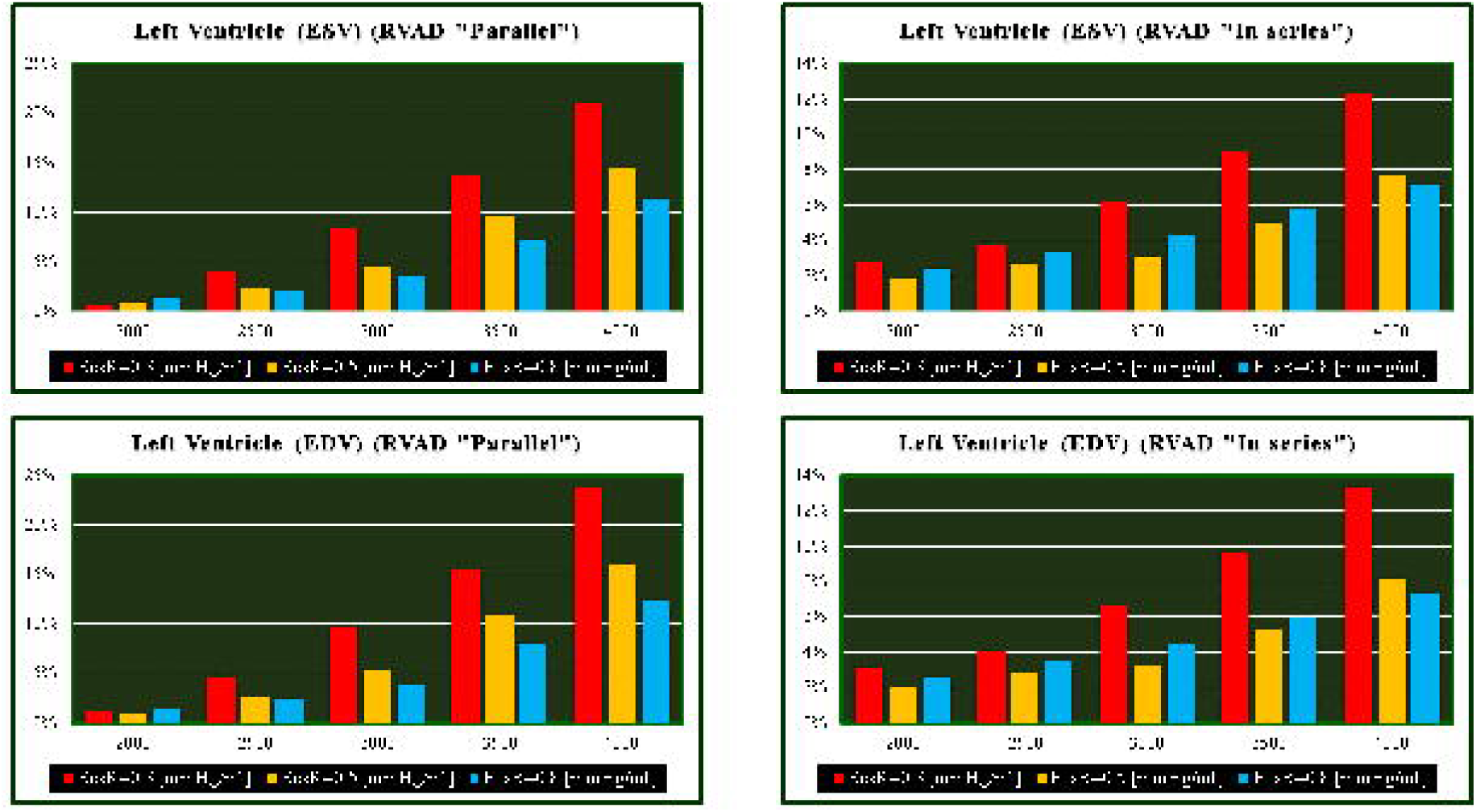
Relative changes calculated in comparison to pathological conditions (EesR_lrl_Ees_RIGHT_=0.3; 0.5 and 0.8 mmHg/ml) for different type of RVAD connection and different rotational speed. For each values of Ees_RIGHT_, the relative change was calculated when the RVAD was connected in parallel and “in series” mode. The left upper (lower) panel shows the relative changes in the left ventricular end systolic (end diastolic) volume when the RVAD was connected in parallel to the right ventricle. The right upper (lower) panel shows the relative changes in the left ventricular end systolic (end diastolic) volume when the RVAD was connected in series to the right ventricle.

**Fig. 6.**
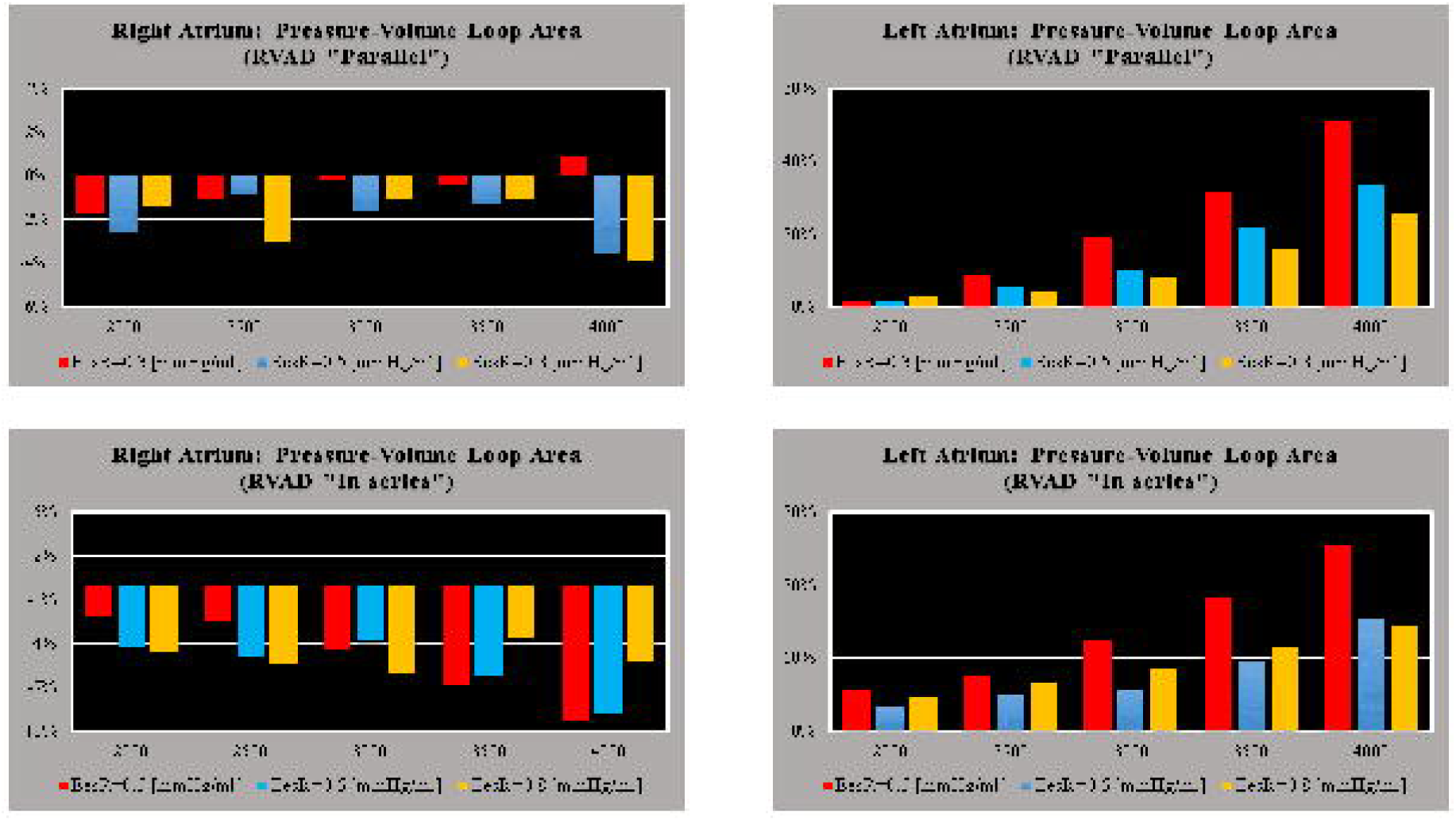
Relative changes calculated in comparison to pathological conditions (EesR_lrl_Ees_RIGHT_=0.3; 0.5 and 0.8 mmHg/ml) for different type of RVAD connection and different rotational speed. For each values of Ees_RIGHT_, the relative change was calculated when the RVAD was connected in parallel and in series mode. The left (right) upper panel shows the relative changes in the right (left) atrium pressure-volume loop area when the RVAD was connected in parallel to the right ventricle. The left (right) lower panel shows the relative changes in the right (left) atrium pressure-volume loop area when the RVAD was connected in series to the right ventricle.

**Fig. 7.**
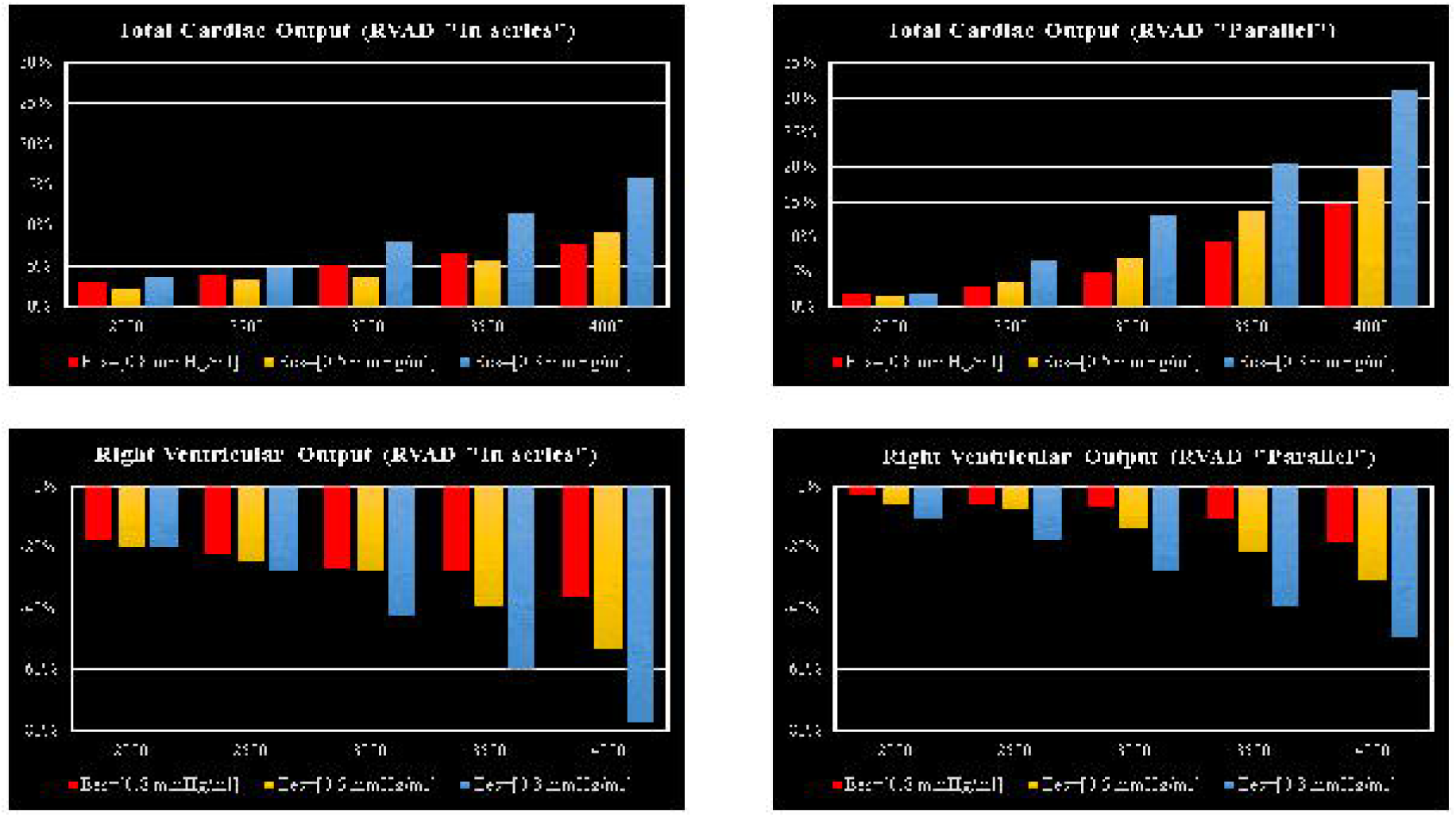
Relative changes calculated in comparison to pathological conditions (EesR_lrl_Ees_RIGHT_=0.3; 0.5 and 0.8 mmHg/ml) for different types of RVAD connection and different rotational speed. For each value of Ees_RIGHT_, the relative change was calculated when the RVAD was connected in parallel and in series mode. The left (right) upper panel shows the relative changes in the total cardiac output (right ventricular flow output plus RVAD flow output) when the RVAD was connected in series (parallel) to the right ventricle. The left (right) lower panel shows the relative changes in the right ventricular flow output when the RVAD was connected in series (parallel) to the right ventricle.

The left upper (lower) panel in Fig. 8 shows the effects induced on left ventricular (atrial) loop when different types of RVAD assistance were applied to pathological conditions (black loops) reproduced by setting Ees_RIGHT_ to 0.3 mmHg/ml. The assistance was applied in parallel (red loops) and in series (blue loops) mode with rotational speed of 3000 rpm.

**Fig. 8.**
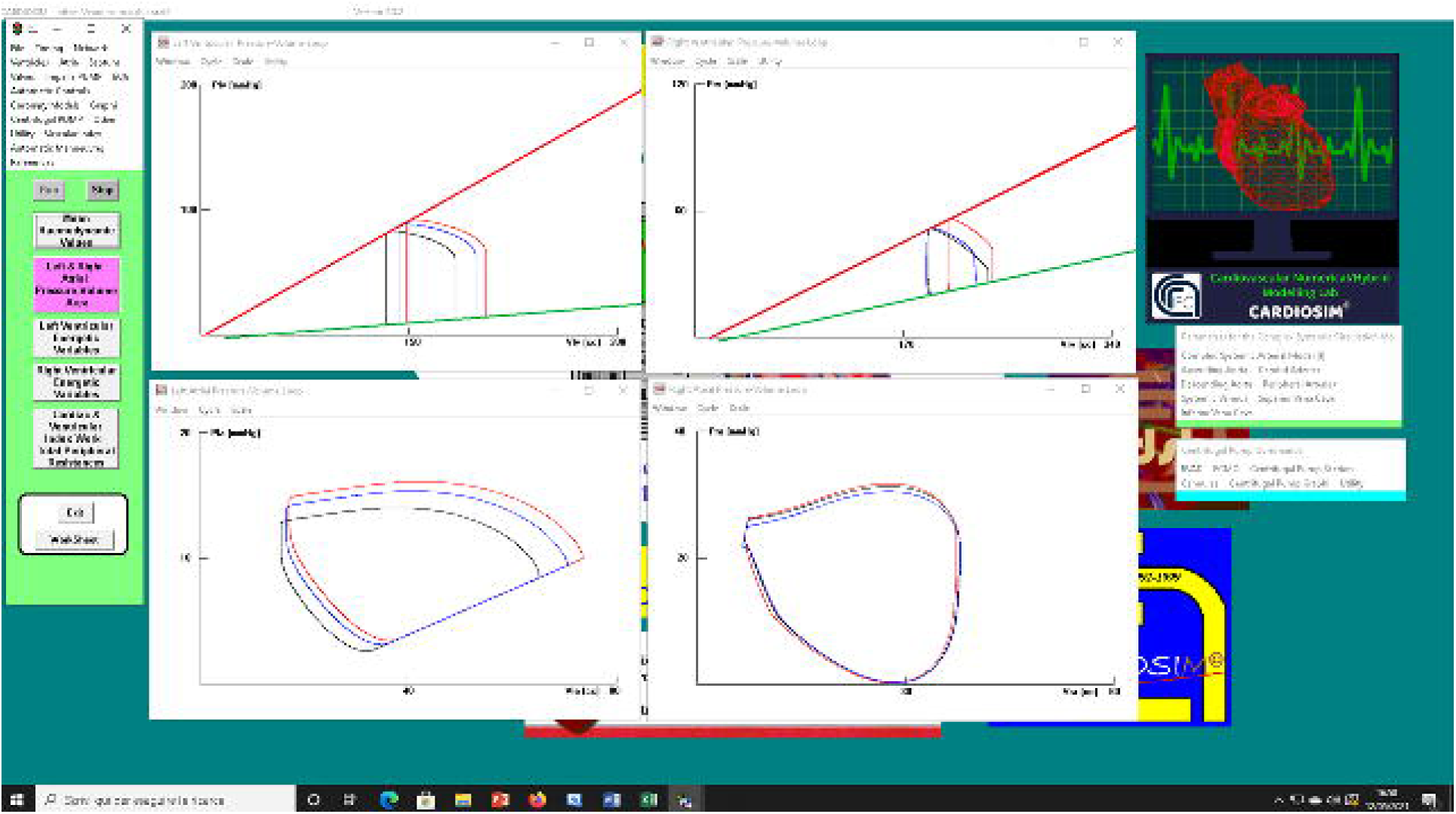
Screen output generated by CARDIOSIM^©^ platform. The left (right) upper panel shows the left (right) ventricular pressure-volume loops when pathological conditions (Ees_RIGHT_=0.3 mmHg/ml) (black line), in-parallel RVAD (pump rotational speed at 3000 rpm) (red line) and in-series RVAD assistance (pump rotational speed at 3000 rpm) (blue line) were reproduced by the simulator, respectively. The left (right) lower panel shows the left (right) atrial pressure-volume loops reproduced by CARDIOSIM^©^.

The right upper (lower) panel shows the effects induced on right ventricular (atrial) loops. When in-parallel assistance was applied, a right-sided shift in the left and right ventricular loop (red lines) was observed. Although in-series RVAD assistance did not cause changes in right end-systolic ventricular volume, it leaded to a reduction in right end-diastolic ventricular loop. The type of assistance did not cause relevant changes in the right atrial loop (right lower panel).

## DISCUSSION

The left ventricle (LV) is coupled to the low-compliance, high-resistance peripheral arterial circulation and is more adaptable to changes in pressure than volume. In contrast, the right ventricle (RV) is coupled to the high-compliance, low-resistance pulmonary circulation and is more adaptable to changes in volume than pressure. The right ventricle consists of a free wall containing a wrap-around circumferential muscle at its base and a septum made of oblique helical fibres crossing each other at 60° angles. This is consistent with the helical ventricular myocardial band concept, which defines two interconnected muscle bands: a basal loop with transverse fibres surrounding the left and right ventricles and an apical loop made of a right- and left-handed helix forming an apical vortex [20],[21]. The wrap-around transverse fibres constrict or compress leading to a bellows motion responsible for 20% of right ventricular output whilst the oblique fibres are responsible for shortening and lengthening, which contribute to 80% of right ventricular systolic function [22]. The crista supra-ventricularis shares muscle fibers with the inter-ventricular septum and the free wall playing a key anatomical and functional role [23]. A reduction in longitudinal contraction and an increase in transverse shortening are observed following cardiopulmonary bypass and pericardiotomy [24]. This is quite an important aspect to bear in mind and may be addressed initially with pulmonary vasodilators [25]. The relationship between structure and function plays a key role in clinical decision-making, which must be based on detailed knowledge of normality and recognise how disease can be addressed to restore normality [22]. The important contribution of right ventricular function has been neglected for a long time due to previous observations and assumptions. The onset of right ventricular dysfunction should trigger the search for the main underlying cause in relation to pressure overload, volume overload or primary myocardial disease [26]. Right heart failure (RHF) is difficult to manage because of its complex geometry and a lack of specific treatments aimed at stabilisation and recovery of right ventricular function. Nevertheless, right ventricular dysfunction remains associated with poor clinical outcome regardless of the underlying disease mechanism [27].

A simulation approach overcomes ethical issues and the risk of offering an ineffective or potentially dangerous therapeutic option. At the same time, it may help focus on the specific problem to address. Our starting point was to develop a failing right heart, which would require support at a subsequent stage. The easier way to do it was to act on the ESPVR slope of both ventricles. A range between 0.35 and 0.74 mmHg/ml is observed in patients with pulmonary hypertension [1] with a cut-off of 0.8 for £eS/£a ratio as the onset of right ventricular maladaptation. Our initial aim was to observe the effects of RVAD support with either in series or parallel connection following stepwise variation of right ventricular end-systolic elastance in patients with increased right ventricular afterload. Therefore, the right ventricular end-systolic elastance considered in the present study ranged between 0.3 and 0.8 mmHg/ml as per previously reported values observed in clinical practice [1],[28],[29]. According to [19], increased native cardiac output is observed in the presence of left ventricular systolic impairment when right ventricular end-systolic elastance increases from 0.1 to 1.0 mmHg/ml. An increased native cardiac output is still observed during VA ECMO support following stepwise increase in right ventricular end-systolic elastance but to a lesser degree. A left-to-right ventricular septal shift is observed during diastole following a stepwise increase in right ventricular end-systolic elastance both with and without VA ECMO support. Our considerations are based in the context of pure RVAD support with either in series or parallel connection.

PVA and EW of the left ventricle gain benefit when the pump speed of the RVAD is at least 3500 rpm. The highest effect is obtained when Ees_RIGHT_ is 0.3 mmHg/ml, which is consistent with significant RV dysfunction (left panels in Fig. 4). The pathological range considered would suggest that early recognition and aggressive RVAD support is advisable where a lower degree of assistance is required generating enough benefit for a less compromised right ventricle with more potential for recovery. This is an important point to consider in the context of LVAD support when the right heart shows signs of failure requiring attention.

The highest beneficial effect is obtained when the RVAD is connected in parallel to the right ventricle with up to nearly 35% increase in total cardiac output (right upper panel in Fig. 7) and lower reduction of right ventricular output compared to in series RVAD connection. No significant effect is observed on the right atrium regardless the type of RVAD connection to the right ventricle. Instead, in-parallel RVAD connection has a more beneficial effect on the left atrium. Again, RVAD support has a more beneficial effect on ESV and EDV of the left ventricle when connected in parallel to the right ventricle (left panels in Fig. 5). The role of the inter-ventricular septum is critical in this context.

Our aim was to observe the effect of pure RVAD assistance at different stages of right ventricular dysfunction to determine appropriate timing for intervention. Our target was early graft failure secondary to right heart dysfunction following orthotopic heart transplant and right ventricular failure following LVAD insertion in an apparently preserved right heart function preoperatively. We have focused our attention mainly on Ees_RIGHT_ neglecting Vo. We have not considered the progressive increase in afterload. Despite these limitation, our preliminary findings support the concept of early intervention in the presence of a failing right heart regardless of its aetiology. This simulation study confirms what had been previously advocated but not always put into practice [30]. A more liberal right ventricular support may be the way forward [31] when considering different support strategies for a failing right ventricle [32]. Late onset of right ventricular failure remains associated with worse survival and higher cumulative incidence of major adverse events [33].

## CONCLUSION

Although RVAD support may be effective in advanced right heart failure, intervention at an earlier stage may require a lesser degree of assistance and be more beneficial for the left ventricle. Early recognition of right ventricular failure followed by aggressive treatment is desirable to achieve a more favourable outcome. RVAD support remains an option for advanced right ventricular failure although the onset of major adverse events may preclude its use. In-parallel RVAD connection to the right ventricle seems a more suitable option.

## Data Availability

All data produced in the present study are available upon reasonable request to the authors

## ACKNOWLEDGEMENTS

None

