## Supplementary material for "Ventricular and Atrial Pressure-Volume Loops: Analysis of the Effects Induced by Right Centrifugal Pump Assistance": Table1

| Table 1 | | |
| --- | --- | --- |
| Symbol | **Description** | **Unit** |
| *P_lv_(t)*  [*P_rv_(t)*] | Instantaneous left (right) ventricular pressure | mmHg |
| *P_lv,0_*  [*P_rv,0_*] | Resting left (right) ventricular pressure | mmHg |
| *V_lv_(t)*  [*V_rv_(t)*] | Instantaneous left (right) ventricular volume | ml |
| *V_lv,0_*  [*V_rv,0_*] | Resting left (right) ventricular volume | ml |
| *e_lv_(t)*  [*e_rv_(t)*] | Left (right) ventricular elastance | mmHg·ml^-1^ |
| *e_Vsp_(t)* | Inter-ventricular septum elastance | mmHg·ml^-1^ |
| *P_la_(t)*  [*P_ra_(t)*] | Instantaneous left (right) atrial pressure | mmHg |
| *P_la,0_*  [*P_ra,0_*] | Resting left (right) atrial pressure | mmHg |
| *V_la_(t)*  [*V_ra_(t)*] | Instantaneous left (right) atrial volume | mL |
| *V_la,0_*  [*V_ra,0_*] | Resting left (right) atrial volume | mL |
| *e_la_(t)*  [*e_ra_(t)*] | Left (right) atrial elastance | mmHg·ml^-1^ |
| *e_Asp_(t)* | Inter-atrial septum elastance | mmHg·ml^-1^ |
