## Supplementary material for "Ventricular and Atrial Pressure-Volume Loops: Analysis of the Effects Induced by Right Centrifugal Pump Assistance": Table2

**Table2	Cardiovascular network**

| *R_pam_ (R_pas_)* | Main (Small) pulmonary arterial resistance [mmHg·cm^-3^·sec] |
| --- | --- |
| *L_pam_ (L_pas_)* | Main (Small) pulmonary arterial inertance [mmHg·cm^-3^·sec^2^] |
| *C_pam_ (C_pas_)* | Main (Small) pulmonary arterial compliance [mmHg^-1^·cm^-3^] |
| *MPAP (SPAP)* | Main (Small) pulmonary arterial pressure [mmHg] |
| *R_par_ (R_pc_)* | Pulmonary arteriole (capillary) resistance [mmHg·cm^-3^·sec] |
| *Wedge* | Pulmonary capillary wedge pressure [mmHg] |
| *C_vp_* | Pulmonary venous compliance [mmHg^-1^·cm^-3^] |
| *R_vp_* | Pulmonary venous resistance [mmHg·cm^-3^·sec] |
| *PVP* | Pulmonary venous pressure [mmHg] |
| *R_ro_ (R_ri_)* | Pulmonary (tricuspid) valve resistance [mmHg·cm^-3^·sec] |
| *SVP* | Systemic veins pressure [mmHg] |
| *C_VS_* | Systemic veins compliance [mmHg^-1^·cm^-3^] |
| *R_VS_* | Systemic veins resistance [mmHg·cm^-3^·sec] |
| *R_Vvs_* | Resistor accounting viscous losses of the  systemic veins wall [mmHg·cm^-3^·sec] |
| *R_SVC_I_* | First Superior vena cava resistance [mmHg·cm^-3^·sec] |
| *C_SVC_* | Superior vena cava compliance [mmHg^-1^·cm^-3^] |
| *L_SVC_* | Superior vena cava inertance [mmHg·cm^-3^·sec^2^] |
| *R_SVC_II_* | Second superior vena cava resistance [mmHg·cm^-3^·sec] |
| *SVCP* | Superior vena cava pressure [mmHg] |
| *LAP(RAP)* | Left (right) atrial pressure [mmHg] |
| *LVP(RVP)* | Left (right) ventricular pressure [mmHg] |
| *R_AA_* | Ascending aorta resistance [mmHg·cm^-3^·sec] |
| *R_Vaa_* | Resistor accounting viscous losses of the  ascending aorta wall [mmHg·cm^-3^·sec] |
| *L_AA_* | Ascending aorta inertance [mmHg·cm^-3^·sec^2^] |
| *C_AA_* | Ascending aorta compliance [mmHg^-1^·cm^-3^] |
| *AoP* | Aortic Pressure [mmHg] |
| *R_DA_* | Descending aorta resistance [mmHg·cm^-3^·sec] |
| *R_Vda_* | Resistor accounting viscous losses of the  descending aorta wall [mmHg·cm^-3^·sec] |
| *L_DA_* | Descending aorta inertance [mmHg·cm^-3^·sec^2^] |
| *C_DA_* | Descending aorta compliance [mmHg^-1^·cm^-3^] |
| *DPA* | Descending aortic pressure [mmHg] |
| *R_CA_* | Carotid arteries resistance [mmHg·cm^-3^·sec] |
| *R_A_* | Peripheral arteries resistance [mmHg·cm^-3^·sec] |
| *R_Va_* | Resistor accounting viscous losses of the  peripheral arteries wall [mmHg·cm^-3^·sec] |
| *C_A_* | Peripheral arteries compliance [mmHg^-1^·cm^-3^] |
| *PA* | Peripheral arteries pressure [mmHg] |
| *R_IVC_, R_IVC_I_, R_IVC_II_* | Inferior vena cava resistances [mmHg·cm^-3^·sec] |
| *L_IVC_* | Inferior vena cava inertance [mmHg·cm^-3^·sec^2^] |
| *C_IVC_* | Inferior vena cava compliance [mmHg^-1^·cm^-3^] |
| *Pt* | Mean intrathoracic pressure [mmHg] |
| *ela (era)* | Left (right) atrial elastance [mmHg/ml] |
| *elv (erv)* | Left (right) ventricular elastance [mmHg/ml] |
| *e_Aspt_ (e_Vspt_)* | Inter-atrial (-ventricular) septal elastance [mmHg/ml] |
| *Q_artery_ (Q_venous_)* | Coronary arterial (venous) flow [ml/sec] |
| *R_oCANN_ (R_iCANN_)* | RVAD output (input) cannula resistance [mmHg·cm^-3^·sec] |
| *L_oCANN_ (L_iCANN_)* | RVAD output (input) cannula inertance [mmHg·cm^-3^·sec^2^] |
| *C_oCANN_ (C_iCANN_)* | RVAD output (input) cannula compliance [mmHg^-1^·cm^-3^] |
| *Qli (Qlo)* | Left ventricular input (output) flow [ml/sec] |
| *Qri (Qro)* | Right ventricular input (output) flow [ml/sec] |
| *Qlia (Qria)* | Left (right) atrial input flow [ml/sec] |
